# Cortical neural state topography reveals mesoscale heterogeneity in autism

**DOI:** 10.1101/2025.07.08.25330892

**Authors:** Daniele Marcotulli, Valentina Francesca Cudia, Martina Vacchetti, Luca Berta, Samuela Morano, Marta Giacobbi, Carlotta Canavese, Barbara Svevi, Federica Silvia Ricci, Federico Amianto, Benedetto Vitiello, Andrea Martinuzzi, Tobias Banaschewski, Argyris Stringaris, Chiara Davico

## Abstract

Characterizing macroscopic brain organization in neuropsychiatric conditions relies on averaging neural activity within discrete regions, yet this approach collapses spatial information that likely carries distinct biological meaning. Here, we introduce a spatial autocorrelation framework to quantify the continuous topographical organization of local states and demonstrate its utility in autism spectrum conditions (ASC). Mapping the spatial heterogeneity of an electroencephalographic (EEG) excitability marker —the aperiodic exponent — across three independent datasets (n = 3767), we show that the autistic cortex exhibits a more heterogeneous topography, as preregistered. This pattern was specific to the mesoscale (∼6–9 cm), replicated across cohorts, and persisted across wakefulness and sleep. This spatial metric outperformed both the global mean and regional variability in predicting ASC status, indicating that topographical arrangement captures biological variance not recovered by conventional approaches. Structural MRI analysis (n = 1198) revealed that local macroanatomy mirrors this functional heterogeneity, with stronger structure–function coupling in ASC, suggesting an anatomical basis for the observed topographical differences. By recovering spatial information typically collapsed through averaging, this framework provides a complementary axis for characterizing macroscopic brain organization across neuropsychiatric conditions.

## Introduction

In any complex biological system, from the synchronization of cardiac cells to the collective motion of animal groups, the spatial arrangement of its constituents determines the system’s dynamic properties. In the human cortex, the smoothness of the excitability landscape, how local neural states are arranged across the cortical surface, regulates how signals propagate across space^1–4^. Here, we test the hypothesis that Autism Spectrum Conditions (ASC), a common neurodevelopmental condition^5^, are characterized by a distinct topological organization, specifically, increased heterogeneity in neural excitability^6^.

Despite its importance for individuals and society, the neurobiological basis of ASC remains elusive^6, 7^. In the search for a neurophysiological mechanism, research has traditionally been divided into two dominant but often disconnected frameworks: the excitation/inhibition (E/I) imbalance hypothesis and the different brain connectivity hypothesis^8–11^, both of which have received substantial independent support but lack integration and failed to provide reliable biomarkers^12^.

The E/I hypothesis suggests that ASC results from altered E/I balance, yet findings remain heterogeneous, likely because studies relied on global averages or isolated regions, obscuring spatial topography^2,13–16^. Similarly, evidence for altered connectivity, such as the “local hyperconnectivity with global hypoconnectivity” hypothesis^12,17,18^ remains vast yet inconsistent.

Here, we argue that these traditionally disconnected frameworks might conceptually converge at the mesoscale, the finest spatial resolution reliably accessible via non-invasive scalp electrophysiology. If local E/I states are heterogeneously distributed across the cortical surface, the resulting spatial gradients could drive a divergent organization of global network dynamics, potentially accounting for findings from both theories within a single spatial mechanism^13,19,20^.

Specifically, we hypothesized that the ASC cortex is characterized by “patches” of relatively distinct excitability states. These patches could arise from region-specific developmental divergences or genetic determinants, reinforcing local connectivity and manifesting as the enhanced local feature focus frequently observed in ASC^11, 17^.

To test this, we quantified the spatial autocorrelation of the aperiodic exponent (slope). While influenced by additional factors including neuromodulatory tone, the aperiodic exponent is sensitive to shifts in excitation-inhibition dynamics^21^ and serves here as a practical electrophysiological index of local neural state. The spatial autocorrelation captures how local neural dynamics at one location correlate with values at other locations, in relation to the distance between electrodes. Therefore, lower spatial autocorrelation indicates the presence of excitability “islands” that contrast sharply with their surroundings (**Fig. 1**). Where local E/I states differ significantly from their neighbors, the resulting gradient would impose a dynamic barrier, constraining the integration between adjacent circuits to follow a divergent trajectory^1,3^.

**Figure 1.**
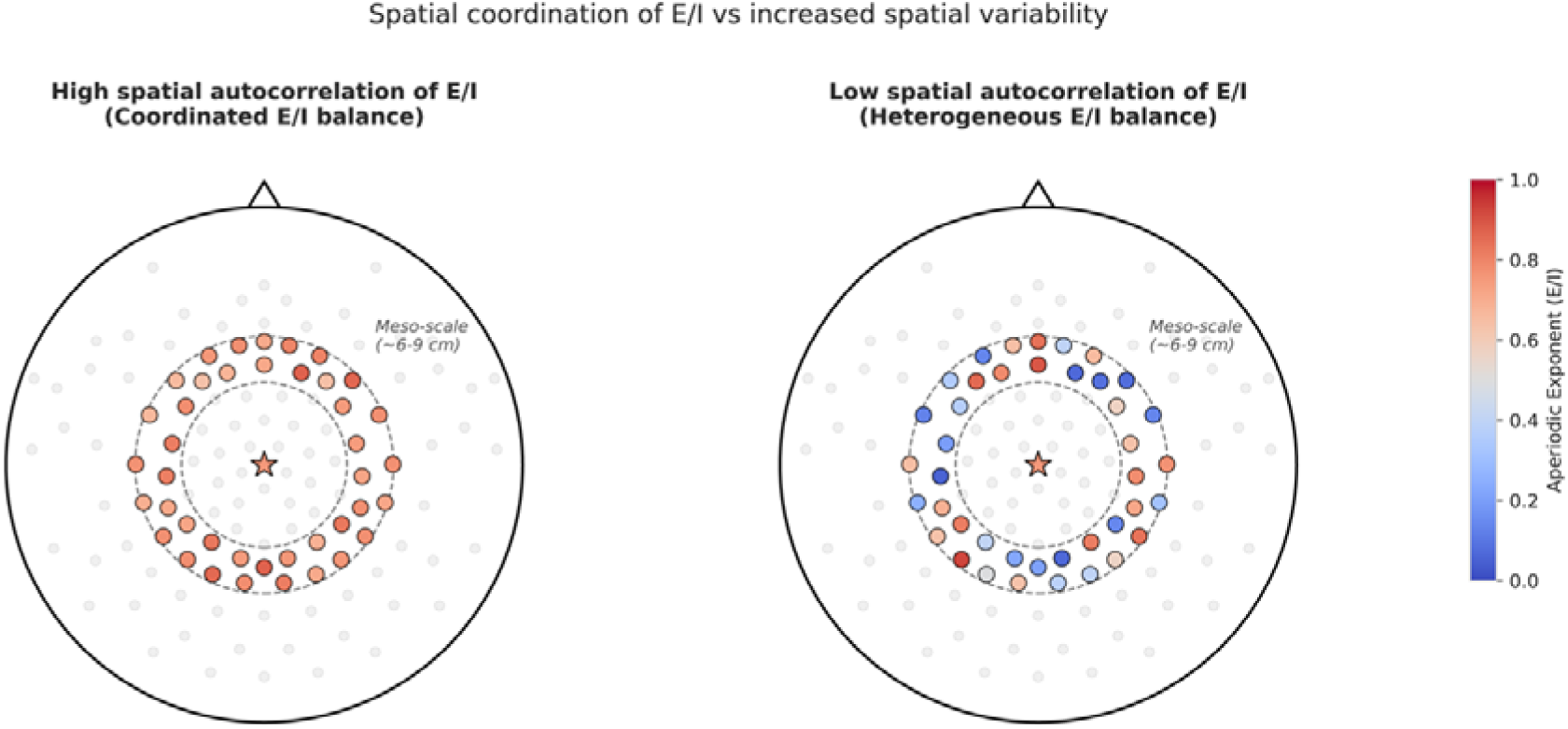
Visual demonstration of increased spatial variability at the mesoscale. Schematic representation of spatial autocorrelation in the aperiodic exponent. The plots illustrate two contrasting spatial organizations of the EEG aperiodic exponent. Each dot (or star) represents a sensor. For a given reference sensor (central star), spatial autocorrelation is calculated based on the similarity of signals within a specific, geometrically defined spatial neighborhood or “lag” (here, the mesoscale band of ∼6–9 cm, denoted by dashed concentric circles). Sensor color reflects the local aperiodic exponent value. (**Left**) A highly coordinated spatial organization (high spatial autocorrelation). Sensors within the mesoscale neighborhood exhibit similar aperiodic exponent values to the reference node and to each other, reflecting a smooth, continuous neural states architecture. (**Right**) A heterogeneous spatial topology (low spatial autocorrelation). Sensors within the exact same mesoscale boundary display highly variable aperiodic values, indicating a “patchy” landscape characterized by abrupt local shifts in neural states.

Crucially, this spatial autocorrelation framework diverges from traditional functional connectivity. Rather than modeling communication between distinct brain regions, our approach maps the continuous spatial distribution of local neural states. In highly complex conditions like ASC, standard regional averaging obscures local excitation/inhibition gradients, washing out crucial neurobiological variance.

By explicitly quantifying spatial heterogeneity rather than averaging it away, Moran’s I transforms cortical distribution into a single metric that still accommodates the possibility that varying neural topographies underlie diverse clinical phenotypes.

To assess whether this heterogeneity is a stable trait rather than a spectral feature driven by sensory processing or active cognition, we extended our analysis to sleep, a state where E/I balance is strictly regulated and external confounds are minimized^22, 23^.

Crucially, if this functional mesoscale heterogeneity is a stable developmental property, it might be plausibly anchored in structural differences at the same spatial scale, offering a multimodal framework to cross-validate these neurophysiological gradients. The local morphological composite (encompassing gray matter, superficial white matter, and their transitional boundary, represented by T1-weighted MRI intensity) served as a natural structural candidate^24–27^.

This study combines pre-registered confirmatory analyses with exploratory extensions. We tested three pre-registered hypotheses:

- The spatial autocorrelation of E/I^21^ is significantly lower in ASC individuals compared to non-autistic controls.
- Increased E/I spatial variability can be observed in autistic individuals also during sleep.
- Greater spatial heterogeneity (i.e., lower spatial autocorrelation) of E/I is associated with more pronounced autism-related traits (Autism Diagnostic Observation Scale-II, ADOS-2).

The pre-registration was conducted in our discovery sample and was later validated in two independent cohorts. See Methods for any deviation.

Additionally, we deployed propensity-score matching to strengthen quasi-causal inferences and triangulated our findings using MRI structure–function correlation. These exploratory (non-pre-registered) analyses, which included the spatial sweep and independent-cohort validation, were designed to assess the robustness and refine the primary hypotheses.

## Results

### The spatial heterogeneity of the aperiodic exponent is greater in ASC individuals compared to non-autistic individuals

In total, 248 autistic subjects were recruited via self-referral or clinician-referral to a specialized tertiary care clinic (**Table 1**). A separate control group of 105 non-autistic individuals was selected from Emergency Room admissions. These patients had no history of epilepsy, acute brain injury, or neurodevelopmental or chronic neurological diagnoses. Clinical characteristics of the sample are presented in **Extended Data Table 1**.

**Table 1.** Demographic characteristics of the sample.

| **Dataset** | **Group** | ***N*, % of total** | ***Age, years, mean***<br>***(SD)*** | ***Males, n***<br>***(%)*** |
| --- | --- | --- | --- | --- |
| **Discovery sample** |  |  |  |  |
|  | **ASC** | 248 (70.3) | 5.5 (3.4) | 196 (79) |
|  | **Non-autistic** | 105 (29.7) | 8.5 (4.8) | 82 (78) |
| **Healthy Brain Network (HBN)<sup>a</sup>** |  |  |  |  |
|  | **ASC** | 416 (16.1) | 10.6 (3.7) | 341 (82.0) |
|  | **ADHD** | 1600 (61.8) | 10.2 (3.3) | 1154 (72.1) |
|  | **Intellectual Disability** | 66 (2.5) | 12.2 (3.5) | 40 (60.6) |
|  | **Non-autistic** | 585 (22.6) | 10.9 (3.9) | 281 (48.0) |
| **NIH** |  |  |  |  |
|  | **AD** | 390 (37.6) | 9.9 (8.1) | 293 (74.9) |
|  | **ASD** | 93 (8.9) | 16.2 (15.1) | 54 (58.1) |
|  | **Non-autistic** | 555 (53.5) | 20.4 (16.6) | 296 (53.3) |

*a. Diagnostic categories are not mutually exclusive*

During wakefulness, autistic individuals exhibited significantly greater spatial heterogeneity of the aperiodic exponent compared to non-autistic individuals (estimate = -0.55, 95% CI: -0.88 to -0.23; ASC n = 130, non-autistic n = 105; **Table 2**). This topographic alteration was independent of age and sex.

**Table 2.** Spatial autocorrelation of the aperiodic exponent during wake across the three datasets.

| **Dataset** | ***Regressors*** | ***Estimates*** | ***95% CI*** | ***p-value*** |
| --- | --- | --- | --- | --- |
| **Discovery** |  |  |  |  |
| **sample** | ASC | -0.55 | -0.88 – -0.23 | **0.001** |
| n = 235 | Sex (M) | -0.24 | -0.63 – 0.14 | 0.207 |
|  | Age | 0.13 | -0.02 – 0.30 | 0.097 |
| **HBN** |  |  |  |  |
| n = 2587 | ASC | -0.34 | -0.62 – -0.06 | **0.016** |
|  | Sex (M) | -0.17 | -0.39 – 0.04 | 0.110 |
|  | Age | 0.25 | 0.15 – 0.35 | **<0.001** |
| **NIH** |  |  |  |  |
| n = 945 | ASC (AD) | -0.25 | -0.43 – -0.08 | **0.005** |
|  | Sex (M) | 0.15 | -0.02 – 0.32 | 0.089 |
|  | Age | 0.12 | 0.04 – 0.21 | **0.005** |

*Bold p-values indicate significance at $\alpha = 0.05$ .*
*Spatial autocorrelation was evaluated at a geodesic distance of 53°.*

During sleep, we also observed a negative association between spatial autocorrelation of the aperiodic exponent and ASC, with an estimate of -0.85 (ASC EEGs n=211; non-autistic EEGs n=94; 95% CI: -1.17 to -0.52, **Extended Data Table 2**). This indicates that the higher spatial heterogeneity in ASC persists across wakefulness and sleep.

The reduced spatial autocorrelation persisted across the following sensitivity analyses: (i) balancing age and sex distributions via optimal propensity-score matching (**Extended Data Table 3**); (ii) excluding participants administered acute sleep aids (melatonin or dimetindene maleate) during recording^28, 29^ (**Extended Data Tables 4a-b**); and (iii) excluding individuals with intellectual disability, global developmental delay, or epileptiform discharges (**Extended Data Tables 5-6**).

### Independent-cohort validation

After testing our primary hypothesis in the discovery cohort, we validated the findings in two independent, publicly available datasets: 1) the Healthy Brain Network (HBN) dataset^30^; 2) a dataset from the National Institute of Mental Health (NIMH) data archive (NIH), previously published by Dede and colleagues^31^.

*1) Healthy Brain Network validation analysis.* In the HBN dataset, we replicated the association between ASC and lower aperiodic exponent spatial autocorrelation against a community-referred non-autistic cohort heavily enriched for ADHD (ASC n = 416, non-autistic n=2171, estimate = -0.34, 95% CI: -0.62 to -0.06, **Tables 1-2**). Similar results were observed after adjusting for other neurodevelopmental conditions and in age and sex matched samples (**Extended Data Table 7a and b**).
*2) NIH datasets validation analysis*^31^. Here too, we found that spatial autocorrelation of aperiodic exponent was lower in the strictly defined autistic (AD) population, in accordance with our hypothesis (**Tables 1-2**) (AD n=390, non-autistic n=555; estimate: -0.25, 95% CI -0.43 to -0.08). This topographic alteration remained significant after accounting for eye closure in a mixed-effects model (**Extended Data Table 8a**). Similarly, when using propensity-score matching for sex and age, we observed an association between autism and higher spatial heterogeneity of the aperiodic exponent (**Extended Data Table 8b**). The association persisted when combining the AD and the lower traits penetrance ASD subgroup (**Extended Data Table 8c**). After propensity score-matching, in this lower trait penetrance subgroup the association did not reach statistical significance (**Extended Data Table 8d**).

### Other spectral features are not consistently more heterogeneous in ASC

Unlike the aperiodic exponent, peak alpha frequency and the aperiodic offset features did not exhibit consistent heterogeneity across datasets. Peak alpha frequency was significantly more heterogeneous in the ASC group within the HBN dataset but failed to replicate both in the discovery sample and in the NIH dataset (**Supplementary Table 1**). Similarly, the aperiodic offset showed increased heterogeneity only in the discovery dataset, and not in HBN (**Supplementary Table 2**).

### ASC is characterized by an increased heterogeneity of the aperiodic exponent specifically in the mesoscale range

To assess the scale specificity of our findings, we systematically mapped the aperiodic exponent variability across a broad spatial range (30°–120°) in the discovery and (20°–120°) HBN dataset. This analysis confirmed that the ASC-associated increase in heterogeneity is not a global phenomenon but predominantly impacts the mesoscale network organization (**Fig. 2A**).

**Figure 2.**
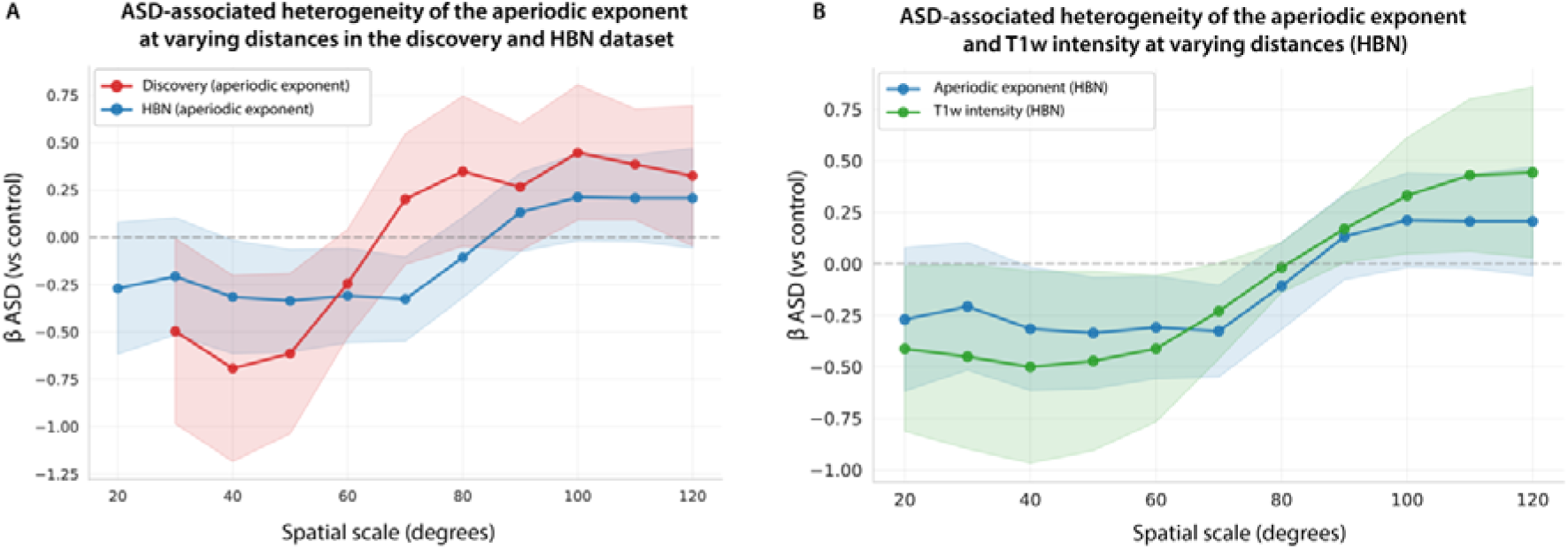
Association between ASC and E/I spatial heterogeneity. (A) Standardized regression coefficients (estimate) for the association between ASC diagnosis and spatial autocorrelation (Moran’s I) of the aperiodic exponent, computed across geodesic distances from 20° to 120°. Negative coefficients indicate greater spatial heterogeneity in the ASC group. Shaded bands represent 95% confidence intervals. The ASC-associated reduction in spatial autocorrelation is concentrated in the mesoscale range (∼42°–64°, corresponding to ∼6–9 cm), converging toward zero at shorter and longer distances. Results are shown as separate lines for the discovery sample (red) and HBN (blue). At short distances (<30° for the 19-channel array), the number of electrode pairs within the kernel bandwidth falls below the threshold for reliable Moran’s I estimation. The NIH dataset was excluded from the scale sweep because its 8 ROIs yield too few spatial pairs per distance band for a stable correlogram. Shaded areas indicate confidence interval. Across spatial lags, the ASD–control difference in spatial autocorrelation showed significant heterogeneity in both datasets, indicating that the effect was scale-specific rather than uniform across distances. In HBN, Cochran’s Q confirmed significant heterogeneity across the 11 tested scales (20°–120°; Q = 37.54, p < 0.001; I² = 73.4%), and meta-regression revealed a significant linear trend toward less negative coefficients with increasing distance (estimate = 0.006 per degree, z = 4.76, p < 0.001), consistent with a group effect that peaks at mesoscale and reverses at longer lags. A similar pattern was observed in the discovery sample across 10 scales (30°–120°; Q = 43.35, p < 0.001; I² = 79.2%; meta-regression estimate = 0.012 per degree, z = 5.90, p < 0.001). Together, these analyses indicate that the ASC-related difference in spatial autocorrelation is not constant across the scalp but is concentrated at mesoscale distances.(B) Standardized regression coefficients for the association between ASC diagnosis and T1w spatial autocorrelation (Moran’s I) across electrode distances (20°–120°). The T1w profile paralleled the EEG pattern: negative at mesoscale distances, returning toward positive values at larger scales. Shaded areas indicate confidence interval.

To test whether mesoscale spatial organization carries diagnostic information beyond the mean or variability of the aperiodic exponent, we compared nested logistic models predicting ASC status (**Extended Data Table 9**). In both cohorts, mesoscale autocorrelation outperformed competing summaries by AIC.

### The mesoscale functional heterogeneity is mirrored at the structural level

Using structural MRI from HBN, we tested whether the spatial profile of T1w intensity mirrors the functional pattern.

The ASC coefficient on T1w spatial autocorrelation across region of interest (ROI) distances showed a profile qualitatively similar to that of the EEG aperiodic exponent: negative at mesoscale distances, returning toward positive values at larger scales (**Fig. 2B**).

A within-subject model revealed a significant EEG × ASC interaction on T1w spatial autocorrelation (estimate = 0.16, CI 0.02 to 0.31), indicating different structure-function coupling in ASC. Specifically, the positive association between functional and structural heterogeneity was stronger in the ASC group. ASC status was also associated with lower T1w spatial autocorrelation (estimate = -0.45, CI -0.86 to -0.03) (**Table 3**). The observed cross-modal coupling remained robust when adjusting for global scan quality, and could not be explained by the mean or variance of gray matter probability within the sampled ROIs (see **Methods**).

**Table 3.** T1w spatial autocorrelation (at 53°) in the HBN dataset.

| **Regressors** | **Estimates** | **95% CI** | **p-value** |
| --- | --- | --- | --- |
| **ASC** | -0.45 | -0.86 – -0.03 | **0.03** |
| **Age** | -0.08 | -0.21 – 0.05 | 0.23 |
| **Sex (M)** | 0.22 | -0.05 – 0.50 | 0.11 |
| **EEG's Moran I** | -0.05 | -0.11 – 0.01 | 0.11 |
| **EEG's Moran I x ASC** | 0.16 | 0.02 – 0.31 | **0.02** |

Subjects with both MRIs and EEGs, n = 1198
*Bold p-values indicate significance at $\alpha = 0.05$ .*
*Spatial autocorrelation was evaluated at a geodesic distance of 53°.*

### Greater spatial heterogeneity of the aperiodic exponent is not associated with more pronounced ASC traits

In the discovery sample, we investigated the association between aperiodic exponent spatial variability and ASC traits in autistic individuals. The ADOS total comparative score had an estimate of 0.00 (EEGs n=184; 95% CI: -0.16 to 0.16), indicating no evidence for the pre-registered hypothesis (**Extended Data Table 10**).

## Discussion

Effective information processing relies on the spatial scale at which local units synchronize. Our findings indicate that in ASC, this arrangement has a divergent organization: the autistic cortex exhibits a more heterogeneous topography at the mesoscale.

While the aperiodic exponent is sensitive to shifts in E/I dynamics, it is not a pure measure of E/I ratio^32^. Nonetheless, our finding, while stemming from the autism E/I hypothesis, does not depend on the exact correspondence between the aperiodic exponent and E/I balance. By mapping the topological organization of the aperiodic exponent across the cortex, we demonstrate that the autistic cortex organizes a well-characterized property of neural activity more heterogeneously across space. This finding was replicated across three independent, large-scale datasets comprising 3767 participants, persisted during sleep, and remained significant after controlling for key confounds such as signal quality, comorbidities, and demographics.

With the above limitations, these results also extend prior reports of altered regional E/I balance in ASC^10,33,34^ by revealing that such imbalances are spatially organized.

This heterogeneity persisted during sleep, indicating a stable, state-independent property not driven by sensory processing or active cognition^22^. Moreover, because Moran’s I was evaluated against spatially permuted nulls, the observed difference reflects spatial organization rather than the value distribution itself.

Volume conduction, which acts as a spatial low-pass filter, is a potential confound^35,36^. However, three lines of evidence suggest that our findings reflect true neural heterogeneity rather than artifacts. First, biophysical simulations confine volume conduction blurring to a radius of 3–4 cm, attenuating rapidly beyond ∼6–7 cm^36^; yet the group difference here is weak at the shortest lags where conduction is strongest, peaks at mesoscale distances and reverses sign beyond ∼65° in the discovery sample and ∼85° in HBN, a profile unlikely to arise from a uniform confound. Second, volume conduction predicts smoother topographies in larger heads; because ASC is associated with macrocephaly in approximately 15% of the population, a head-size artifact would theoretically result in increased spatial autocorrelation. Our observation of the opposite pattern (increased spatial heterogeneity) indicates that the neural signal dominates the physical conduction effect. Third, a generalized conduction artifact would uniformly affect all spectral features, yet our findings were metric-specific: while the aperiodic exponent exhibited robust spatial heterogeneity, the aperiodic offset showed no such effect (e.g., in the HBN dataset, the ASC coefficient for offset heterogeneity was null: estimate = -0.02, p = 0.88).

The spatial specificity of our findings, peaking at the mesoscale (∼6–9 cm), may provide novel insight into the nature of E/I dysregulation in ASC. This window exceeds the radius of maximum passive volume conduction (∼3–4 cm), making a simple signal-leakage account unlikely, yet remains local enough to capture the integration of distinct functional territories before global signal dominance takes over. The increased heterogeneity at this specific scale suggests that the neurobiological deficit in ASC may not be a failure of local micro-scale circuits nor a whole-brain state shift at the macro-scale, but rather an alteration in the functional coordination of neighboring neural populations. Formal model comparison reinforced this interpretation: mesoscale autocorrelation outperformed both the channel-wise mean and standard deviation of the aperiodic exponent in predicting ASC status. The diagnostic information carried by Moran’s I thus reflects the spatial arrangement of excitability states, not their central tendency or dispersion, ruling out the possibility that the spatial signal merely recapitulates increased scatter across channels or average shifts in neural states.

The relevance and specificity of the mesoscale range was further evidenced at the structural level, where we observed regional heterogeneity in bias-corrected T1w signal intensity. At the ROI dimensions utilized, the T1w signal reflects a local morphological composite, capturing regional folding geometry, superficial white matter, and cytoarchitectural tissue organization^37^. While this composite extends beyond the synaptic architecture generating the EEG signal, it serves as a macroscopic proxy for local neuroanatomy. The coupling between structural and functional heterogeneity was significantly stronger in autistic individuals, suggesting that these two dimensions share a spatial organization in ASC. Consistent with this, Stoner et al. identified patch-like cortical architecture in autistic individuals^38^.

The association between reduced spatial autocorrelation and ASC was replicated across all three datasets. However, in the NIH-derived dataset the association was absent when combining the autism disorder subgroup with broader ASC criteria. The attenuation pattern observed in this dataset is consistent with a signal that is modulated by both diagnostic stringency and spatial measurement resolution: the NIH dataset’s coarse 8-ROI parcellation may lack the granularity to resolve mesoscale structure. Furthermore, recent evidence suggests that as diagnostic criteria for neurodevelopmental conditions expand, the underlying genetic and biological coherence tends to decrease^39^. Accordingly, the inclusion of older individuals with lower trait penetrance in the NIH cohort likely introduces a broader, more heterogeneous biological landscape, blurring the distinct mesoscale E/I topography that specifically characterizes the more stringently defined neurodevelopmental phenotype in our primary analysis.

However, within the ASC group, aperiodic exponent spatial autocorrelation was not associated with autism trait penetrance as indexed by the ADOS comparative total score. The absence of association with ADOS score, combined with the robust neurobiological signal, suggests that mesoscale E/I heterogeneity may index a dimension of neural organization that does not map into current summative behavioral phenotyping^40^.

Our results remained consistent even when controlling for co-occurring neurodevelopmental diagnoses (such as ADHD and intellectual disability), supporting the robustness of this signal within our ASC cohort. However, whether this mesoscale spatial decoupling is uniquely specific to ASC or represents a more general neurodevelopmental feature remains an open question. Much like the extensive genetic pleiotropy observed across psychiatric boundaries^41^, this topographic E/I signature may reflect a transdiagnostic biological vulnerability. This potential pleiotropy could partly explain why the spatial signal does not scale linearly with ASC-specific behavioral metrics (e.g., the ADOS); rather, this neurophysiological consistency coexists with marked clinical heterogeneity, underscoring that a shared biological feature does not dictate a singular functional outcome.

Future studies with more refined clinical characterization will be essential. The spatial distribution of EEG spectral features could help cluster autistic individuals into phenotypically meaningful subtypes^41^, and mapping individual E/I topography could inform spatially targeted neuromodulation.

More broadly, the analytic framework developed here, i.e. mapping the spatial autocorrelation of spectral parameters across the cortical surface, is not specific to autism. E/I dysregulation features in schizophrenia, ADHD, and epilepsy, yet these conditions are typically studied through approaches blind to spatial topology. Mesoscale spatial organization of neural excitability may represent a general principle whose divergence characterizes multiple neurodevelopmental conditions. Notably, this can be detected using scalp EEG, making this insight accessible through common clinical tools.

These results should be interpreted in light of a few limitations. First, in all evaluated cohorts, ASC diagnoses and assessments were performed at multiple centers and by different clinicians. While this introduces noise, the persistence of the E/I signature across these unstandardized environments suggests a robust phenotype relevant to real-world practice. Indeed, the effect was largest in the discovery sample, where the clinical control group may have contributed to a more conservative baseline and attenuated but significant in both community-recruited cohorts. Second, potential referral biases, unequal phenotypic characteristics across the diverse cohorts, and incomplete clinical characterization represent inherent limitations of our study design, which we attempted to account for through targeted sensitivity analyses.

About mesoscale specificity, scalp EEG provides more reliable spatial estimates at mesoscale and longer distances than at shorter ones, where fewer electrode pairs and greater positioning uncertainty reduce statistical power. As a result, our correlogram can more confidently distinguish the mesoscale effect from longer-distance patterns than from shorter-distance ones.

Regarding MRI, while our ROIs effectively capture local mesoscale morphological composites, they inherently encompass tissue boundaries and partial volume effects, precluding strict layer-specific claims about pure intracortical myelination. Moreover, our radial projection method identifies the anatomical center of each sensor’s macroscopic lead field, representing a heuristic approximation that cannot fully account for the spatial smearing inherent to EEG volume conduction.

This study identifies spatially heterogeneous neural dynamics as a fundamental neurobiological feature of ASC. By shifting focus from global averages to mesoscale topology, our results indicate that the spatial arrangement of local neural states carries mechanistic information. The analytic framework developed here is generalizable beyond autism and beyond E/I: mapping the spatial autocorrelation of any neural parameter across the cortical surface offers a scalable, spatially informed approach to characterize macroscopic brain organization using clinical EEG.

## Materials and methods

### Discovery Sample’s Participants

This retrospective study was based on clinical data collected from participants who attended the neurological Day Hospital at the Regina Margherita Children’s Hospital, University of Turin, between January 30, 2018, and May 29, 2024.

Inclusion criteria for the case group were the following:

- Children (age<18 years) with a previous diagnosis of autism spectrum disorder made by an expert child and adolescent neuropsychiatrist, using standard diagnostic instruments.
- Written informed consent for research purposes signed by parents or legal guardians.
- An EEG performed on the date of the admission.

To ensure our sample was representative of the broad clinical population, we adopted an inclusive recruitment strategy without restrictive exclusion criteria. All individuals with a confirmed diagnosis of ASC who attended the neurological day hospital and completed an EEG recording were included in the analysis, preserving the natural heterogeneity of the cohort.

The control group for this study was derived from hospital data of participants (children under 18 years of age) admitted to the emergency department during the same period. These participants were referred for neuropsychiatric evaluation due to symptoms such as headache, loss of consciousness, alteration of consciousness or paroxysmal movements. Exclusion criteria applied to the control group included a diagnosis of developmental disorder, epilepsy, acute brain injury, or seizure.

### 248 participants were recruited in the autistic group, and 105 participants were recruited in the non-autistic group

All the procedures were approved by the Citta della Salute e delle Scienza ethics committee and parents or legal tutors of ASC individuals gave informed consent. Control EEGs were collected anonymously.

### Publicly available datasets

#### Healthy Brain Network

EEG data and participant demographics from the Healthy Brain Network (HBN) (Releases 1–11 and the NC Release with available BIDS-formatted data) were obtained from the public repository ( https://fcon_1000.projects.nitrc.org/indi/cmi_healthy_brain_network/MRI_EEG.html). The HBN uses a community-referred recruitment model in the New York City area and collects, stores, and openly shares de-identified data on psychiatric, behavioral, cognitive, and lifestyle phenotypes (e.g., fitness, diet), along with multimodal brain imaging (MRI), electroencephalography (EEG), genetics, and actigraphy for children and adolescents aged 5–21 years. EEG acquisition protocols and diagnostic criteria for the HBN cohort are described elsewhere^30^.

#### Sample collected from Dede et al. (NIH)

The sample combined by Dede and colleagues^31^ and derived from the National Institute of Mental Health (NIMH) data archive (https://nda.nih.gov/edit_collection.html?id=2022) were obtained from https://github.com/adede1988/SheffieldAutismBiomarkers. In this dataset, we used the aperiodic exponent normalized to the power spectrum distribution as our E/I measure to mitigate inter-dataset differences in overall signal amplitude. Since our hypothesis focused on autism, we included the “autistic disorder” (AD) group (using the original datasets definition), the lower trait penetrance according to the ADOS nomenclature “ASD” subgroup^31^ and the non-autistic controls, excluding the schizophrenia and bipolar cohorts from the Bipolar and Schizophrenia Consortium for parsing intermediate phenotypes (**Table 1**). We first isolated the strictly defined AD group to test for spatial E/I divergence in the cohort with the most pronounced trait penetrance. We then analyzed the combined AD and broader “ASD” cohorts against controls to test whether the topographic heterogeneity generalizes across the wider diagnostic spectrum.

Both the HBN and the NIH-derived cohorts provided only wakefulness EEGs. In the NIH dataset, we prioritized eyes-open recordings to maximize sample size.

### Measures and EEG Recordings in the discovery sample

Demographic and clinical data were collected through electronic medical records. For the case group, the collected data included: age at the admission, sex, comorbidities, traits penetrance based on ADOS score, and EEG description. For the control group, the collected data included: age, reason of admission, sex, anamnestic data, comorbidities, EEG description.

According to clinical and technical considerations, EEGs were recorded during wakefulness, sleep or both states.

The sleep recordings also served to exclude that increased E/I variability is associated with information and/or sensory processing that occurs during wakefulness, or wake-related artifacts (e.g. muscle activity).

Children in the case group underwent baseline standard EEG recording as part of a comprehensive diagnostic protocol. For children in the control group, EEG recordings were performed during their diagnostic evaluation for reasons other than neurodevelopmental conditions, epilepsy and acute brain injury.

Trained pediatric neurologists evaluated the EEGs for presence and localization of epileptiform discharges.

### EEG Data Analysis

#### Extraction of E/I measure

To derive E/I from EEG segments, we employed the spectral parameterization technique described by Donoghue and colleagues^21^. We focused on the exponent of the aperiodic component, a well characterized electrophysiological proxy for E/I balance. Power spectral density (PSD) was computed using Welch’s method (1–40 Hz range). We then employed the spectral parameterization algorithm (*specparam*, formerly *FOOOF*) to decompose the PSD into periodic oscillatory peaks and an aperiodic 1/f exponent background.

For the NIH dataset we used data already available at https://github.com/adede1988/SheffieldAutismBiomarkers.

For the NIH dataset, in which each ROI represents an average across multiple electrodes from heterogeneous recording setups, we used the aperiodic exponent normalized to the power spectrum distribution to mitigate inter-study amplitude biases.

#### Moran’s I extraction

To quantify the spatial organization of the E/I landscape, we computed Moran’s I for the aperiodic exponent across the scalp surface.

Spatial autocorrelation was assessed using geodesic metrics to account for cranial topology and arc lengths (great-circle distances) on a unit sphere, targeting a critical mesoscale window of ∼42–64° (corresponding to ∼6–9 cm on a 51 cm circumference cranium). This scale exceeds the radius of maximum passive volume conduction (∼3–4 cm) to capture genuine intra-hemispheric functional coordination (e.g., fronto-parietal integration) rather than conductivity artifacts. We applied an inverse-distance kernel with a constant weighted-mean neighbor angle of ∼53° (∼7.5 cm) to emphasize mid-range interactions. To ensure consistent graph connectivity across varying resolutions, bandwidths were adapted inversely to electrode density (8° for 128-channel, 11° for 19-channel, and 14° for 8-ROI datasets). The primary target distance of 53° was selected during analysis development to center the kernel above the dominant volume conduction radius and centered on the mesoscale; the subsequent scale sweep (30°–120° for the discovery sample; 20°–120° for the higher resolution HBN dataset) confirmed that the effect is not contingent on this specific choice. Because geodesic angles are scale-invariant, the spatial weights are unaffected by head size variation across the age range; the centimeter equivalents cited throughout are approximate, based on a representative 51 cm head circumference (50th percentile for a 6-year-old boy, close to the mean age of the discovery sample).

To verify that our results were not an artifact of the chosen spatial weighting scheme, we conducted a sensitivity analysis specifically utilizing the higher-resolution HBN dataset (128 electrodes). In addition to the primary inversely weighted kernel, we re-computed the spatial autocorrelations for both the EEG and MRI data using a continuous Gaussian spatial weight matrix. The weight w_ij_ between electrodes I and j was defined as:

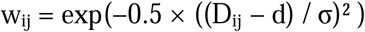

where D_ij_ is the angular distance (in degrees), d is the target center distance of the spatial filter, and σ is the bandwidth (fixed at 16°, corresponding to an approximate cortical distance of 23 mm).

We applied this kernel to the HBN dataset only because a similar finely tuned spatial filtering is methodologically robust only for high-density arrays (e.g., 128 channels), where the dense spatial sampling provides sufficient pairs at varying distances.

To ensure consistent graph connectivity across the vastly different spatial resolutions of the three datasets, we adapted the kernel bandwidth (δ) inversely to electrode density:

- HBN (128-ch): Bandwidth = ±8°.
- Discovery sample (19-ch): Bandwidth = ±11°.
- NIH (8 ROIs): Bandwidth = ±14°.

Consequently, coarser arrays needed to integrate over a wider spatial band, which may dilute scale specificity. Furthermore, for the HBN array, spatial weights were computed using a regularized inverse-distance kernel with an offset = 5°. This offset corresponds to the order-of-magnitude positional uncertainty inherent to high-density electrode caps: cap fit variability and inter-individual head geometry differences introduce electrode displacement errors of approximately 3–7° in geodesic angle. Below this threshold, nominal distance differences carry no reliable spatial information. The offset therefore acts as a nugget correction that prevents the weight function from making spurious distinctions between pairs whose geodesic separations differ by less than the measurement precision. For the 19-channel and 8-ROI arrays, the wider kernel bandwidths (±11° and ±14°) already incorporate this positional uncertainty, making the offset unnecessary.

This adaptive approach ensured that the spatial graph remained connected (reducing the probability of isolated nodes) while targeting the same physical mesoscale distance in all cohorts. Weights were row-standardized (“R” transform). Moran’s I was computed against a reference distribution generated via 499 random permutations. Lower (negative) Moran’s I values indicate greater spatial heterogeneity.

The spatial sweep analysis (30°–120° in the discovery sample; 20°–120° in the HBN) was designed to characterize the distance-dependent profile of the ASC-E/I association, not to test significance at each individual distance. We therefore report the full coefficient curve with 95% confidence intervals across all distances and interpret the overall shape of the profile rather than applying pointwise significance thresholds. This approach avoids the inferential distortions of multiple pointwise tests while preserving sensitivity to the scale-specificity of the effect.

### T1-weighted MRI Preprocessing Electrode-matched MRI extraction

Structural MRI data were obtained from the Healthy Brain Network (HBN) biobank. We utilized the preprocessed T1-weighted (T1w) volumes generated by the Configurable Pipeline for the Analysis of Connectomes (C-PAC)^42^. These data, available via the AWS Open Data Registry (s3://fcp-indi/data/Projects/HBN/CPAC_preprocessed/), were processed using standard anatomical registration workflows, resulting in skull-stripped, N4 bias-corrected images non-linearly registered to the MNI152 template space at 2 mm isotropic resolution (transform_Warped.nii.gz). To map functional EEG sensors to underlying cortical structure, we utilized the standard GSN-HydroCel-129 montage geometry (EGI, Eugene, OR). Electrode positions were mathematically transformed from the MNE-Python “Head” coordinate frame (Neuromag convention: +X= Nasion, +Y= Left, +Z= Superior) to the MNI152 RAS coordinate system (+X= Right, +Y= Anterior, +Z= Superior) via a 90-degree axial rotation. For each electrode, a search vector was defined originating from the montage center (0,0,0) and extending radially outward. We sampled the MNI152 2mm Gray Matter (GM) probability template along this vector at depths ranging from 55 to 85 mm (2 mm steps). The cortical coordinate for each electrode was defined as the depth maximizing GM probability. To ensure valid cortical sampling, electrodes projecting to non-cortical tissue (e.g., orbits, jaw, sinuses) were identified and excluded. Electrodes were retained only if their projected 3 mm sphere captured a mean GM probability ≥ 0.30 (or ≥ 0.50 in a sensitivity analysis). This procedure resulted in a final set of 119 validated (117 in the sensitivity analysis with more stringent GM probability inclusion criteria) cortical electrodes for downstream analysis.

Structural morphology was quantified by extracting T1w intensity values at each validated cortical coordinate. For every subject (n = 1198), we defined spherical Regions of Interest (ROIs) centered on the 119 (117 in the sensitivity analyses) projected coordinates. The primary analysis utilized a 4 mm radius sphere, selected to capture the mesoscale “morphological composite” (local gyrification and tissue contrast) of a single cortical gyrus while minimizing overlap between adjacent sensors. Sensitivity analyses were performed using radii of 3 mm and 6 mm. The mean T1w intensity was calculated within each sphere using Python’s nilearn. While raw T1w intensities are typically arbitrary units dependent on scanner gain, our downstream use of Moran’s I (a ratio of covariances) renders the analysis scale-invariant, effectively normalizing global intensity differences between subjects.

Coordinate validation confirmed no systematic between-group bias in ROI placement, as neither the mean nor the coefficient of variation of GM probability within the spherical ROIs differed by diagnosis. Furthermore, the cross-modal dissociation test yielded a null interaction (EEG_Moran_I x ASC) when evaluating GM probability (p > 0.05), suggesting that the topological decoupling observed in the ASC group is specifically driven by T1w signal rather than macroscopic differences in partial-volume tissue fractions.

While the spherical volumetric sampling offers lower cytoarchitectural specificity and gyral homology than surface-based parcellations, it was specifically chosen to accommodate the spatial constraints of the EEG array. To accurately compare spatial autocorrelation across modalities, it is mathematically required that the structural and functional spatial weight matrices (W) are identical. Employing spherical ROIs based on electrode projections guarantees the topological equivalence.

Furthermore, volume conductor models demonstrate that EEG scalp signals are influenced by the complex geometry of cortical folding and the distinct conductivity of the underlying white matter.

### Statistical Analysis

To test the hypothesis that greater spatial heterogeneity in the E/I balance is associated with ASC features, we used Moran’s I to quantify spatial heterogeneity and the aperiodic exponent as an electrophysiological proxy for E/I^21,32,43^. Moran’s I is a measure of spatial variability approximately centered on zero, such that positive values indicate clustering and negative values indicate dispersion (**Fig. 1**).

For analyses spanning multiple behavioral states (sleep, resting-state), we fit linear mixed-effects models with subject-specific random intercepts. Associations between clinical variables and E/I Moran’s I, where no grouping structure was required, were tested with linear regression. All continuous variables were standardized. In matched samples, matching weights were used as regression weights.

Within-subject EEG–MRI spatial coupling in ASC and non-ASC participants was tested using the linear model:

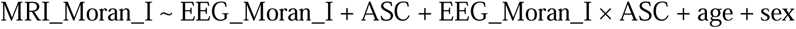

where EEG_Moran_I is the electrode-level Moran’s I of the aperiodic exponent (eyes-closed), and MRI_Moran_I is the T1w Moran’s I at a given distance. The EEG_Moran_I × ASC interaction term tests whether the strength of structure–function spatial coupling differs between groups. We swept the MRI distance from 20° to 120° in 10° steps to assess the shape of the association across spatial scales.

To verify robustness against potential confounds, we performed parallel control analyses including: (i) T1w image quality metrics (coefficient of variation, entropy), (ii) head motion (mean framewise displacement), and (iii) site-specific scanner performance metrics (from MRIQC), demonstrating that the observed effects were not driven by data quality or acquisition artifacts.

All reported p-values are two-sided.

Claude Code (Anthropic, Claude Opus), integrated in the Visual Studio Code IDE, was used to assist with code linting and commenting. The authors reviewed all outputs and take full responsibility for the accuracy and integrity of all code, analyses, and results.

### Study registration, open science and code availability

The three main hypotheses and their analyses were registered on the Open Science Framework before analysing the data (https://osf.io/4q83m). Python codes for data cleaning and analysis are available in GitHub at https://github.com/danielemarcotulli/mesoscale_architecture_autism.git.

### Deviation from the pre-registered analyses

While the study adhered to the core pre-registered protocol (https://osf.io/4q83m/) the following deviations were made. First, we expanded the spatial analysis from a single distance to a sweep across 30°–120° / 20°–120° to characterize scale-specificity. Second, we added propensity-score weighting and medication exclusions as robustness checks. Third, we extended validation to two independent datasets (HBN and NIH). Fourth, we analyzed MRIs from the HBN dataset to evaluate structure-function coupling. These analyses are exploratory and are distinguished from confirmatory results throughout.

## Acknowledgments

We thank the UCL MEG group and Dr. Daniel Rautio for offering constructive feedback and discussion. Claude Code (Anthropic, Claude Opus) was used for code linting and commenting. The authors take full responsibility for the accuracy and integrity of all code, analyses, and results.

## Funding

This work received no specific funding.

## Author contributions

Conceptualization: DM

Methodology: DM

Software: DM

Investigation: DM, VFC

Data Curation: VFC, DM

Validation: DM

Formal analysis: DM

Resources: CD, MV, BS, MG, SM, FSR, CC

Visualization: DM, LB

Project administration: DM

Supervision: AS, CD

Writing – original draft: DM, VFC

Writing – review & editing: All Authors

## Competing Interests

In the last 2 years, DM has received consultant honoraria from Angelini (Ethos).

In the last 2 years, BV has received consultant honoraria from Simon-Kucher, Medice, Angelini (Ethos), and Alkermes Pharmaceuticals.

In the last 2 years, TB has received consultant honoraria from AGB pharma, Infectopharm, Medice, Neuraxpharm, Neurim Pharmaceuticals, Oberberg GmbH and Takeda, and speaker’s fee by AGB pharma, Janssen-Cilag, Medice, Oberberg GmbH and Takeda.

## Data availability

Data used for results validation from the dataset of Dede et al.^31^ are available at https://github.com/adede1988/SheffieldAutismBiomarkers.

Use, sharing, adaptation, distribution, and reproduction are permitted under the Creative Commons Attribution 4.0 International License (CC BY 4.0), provided appropriate credit is given to the original author(s) and the source, a link to the license is provided, and any changes are indicated. Images or other third-party material are included under the article’s Creative Commons license unless otherwise stated; for uses not covered by the license or permitted by law, permission may be required from the rights holder. License: http://creativecommons.org/licenses/by/4.0/.

EEG data from the Healthy Brain Network (HBN) are freely available from the HBN portal: https://fcon_1000.projects.nitrc.org/indi/cmi_healthy_brain_network/. Access to associated phenotypic data is granted upon request via the same website, following the procedures described there.

Discovery sample’s individual-level EEG and clinical data contain protected health information from minors and cannot be made openly available under the terms of the ethical approval granted by the Citta della Salute e delle Scienza ethics committee. Fully de-identified data are available from the corresponding author upon reasonable request. Access requests will be reviewed within 6 weeks and granted subject to a data sharing agreement compliant with applicable privacy regulations (EU General Data Protection Regulation).

## EXTENDED DATA

**Extended Data Table 1.** Demographic, clinical and EEG-related characteristics of the discovery cohort.

| Variable | ASC (N=248) | Non-autistic (n=105) |
| --- | --- | --- |
| Sex, n (% males) | 196 (79) | 82 (78) |
| Age, months, mean (SD) | 66.1 (41.3) | 101.5 (57.2) |
| Age range, months | 20-202 | 15-215 |
| Intellectual Disability, n (%) | 155 (62) | 0 (0) |
| Comorbidities, n (%) | 88 (35) | 31 (29.5) |
| EEG anomalies reported, % | 153 (61.7) | 48 (45.7) |
| EEG state (n) |  |  |
| Wake | 130 | 105 |
| Sleep | 211 | 94 |
| Medication during EEG (n) |  |  |
| Melatonin | 152 | 53 |
| Dimetindene Maleate | 60 | - |
| Reason for admission non-autistic only, n (%) |  |  |
| Loss of consciousness | - | 38 (36.2) |
| **Non-epileptic paroxysmal events** | - | 28 (26.7) |
| **Headache** | - | 25 (23.8) |
| **Altered consciousness** | - | 6 (5.7) |
| **Vertigo** | - | 6 (5.7) |
| **Other** | - | 2 (1.9) |

**Extended Data Table 2.** Aperiodic exponent (slope) spatial autocorrelation during sleep in the discovery cohort.

| **Regressor** | **Estimate** | **95% CI** | **p-value** |
| --- | --- | --- | --- |
| **ASC** | -0.85 | -1.17 – -0.52 | **<0.001** |
| **Age** | 0.12 | -0.04 – 0.27 | 0.139 |
| **Sex** | -0.09 | -0.46 – 0.28 | 0.630 |

EEGs n = 305
*Bold p-values indicate significance at $\alpha = 0.05$ .*
*Spatial autocorrelation was evaluated at a geodesic distance of 53°.*

**Extended Data Table 3.** Aperiodic exponent spatial autocorrelation during wake in the age and sex matched samples (discovery cohort).

| **Regressor** | **Estimate** | **95% CI** | **p** |
| --- | --- | --- | --- |
| **ASC** | -0.62 | -0.96 – -0.29 | **<0.001** |
| **Age** | 0.14 | -0.03 – 0.31 | 0.098 |
| **Sex** | 0.28 | -0.13 – 0.69 | 0.175 |

EEGs n = 210
*Bold p-values indicate significance at $\alpha = 0.05$ .*
*Spatial autocorrelation was evaluated at a geodesic distance of 53°.*

**Extended Data Table 4a.**
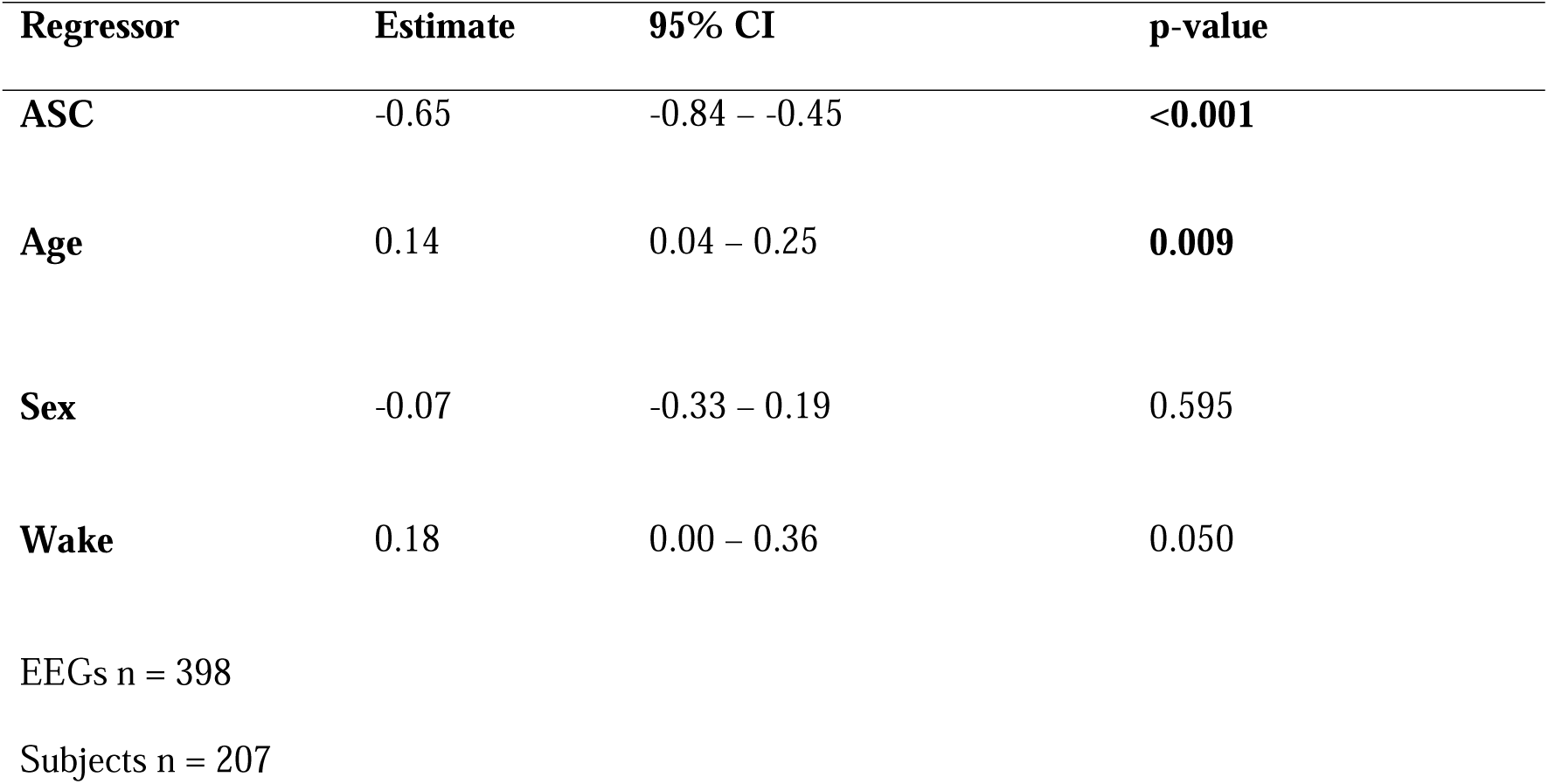
Aperiodic exponent spatial autocorrelation excluding participants who received dimetindene maleate in both sleep and wakefulness EEG (discovery sample).

**Extended Data Table 4b.**
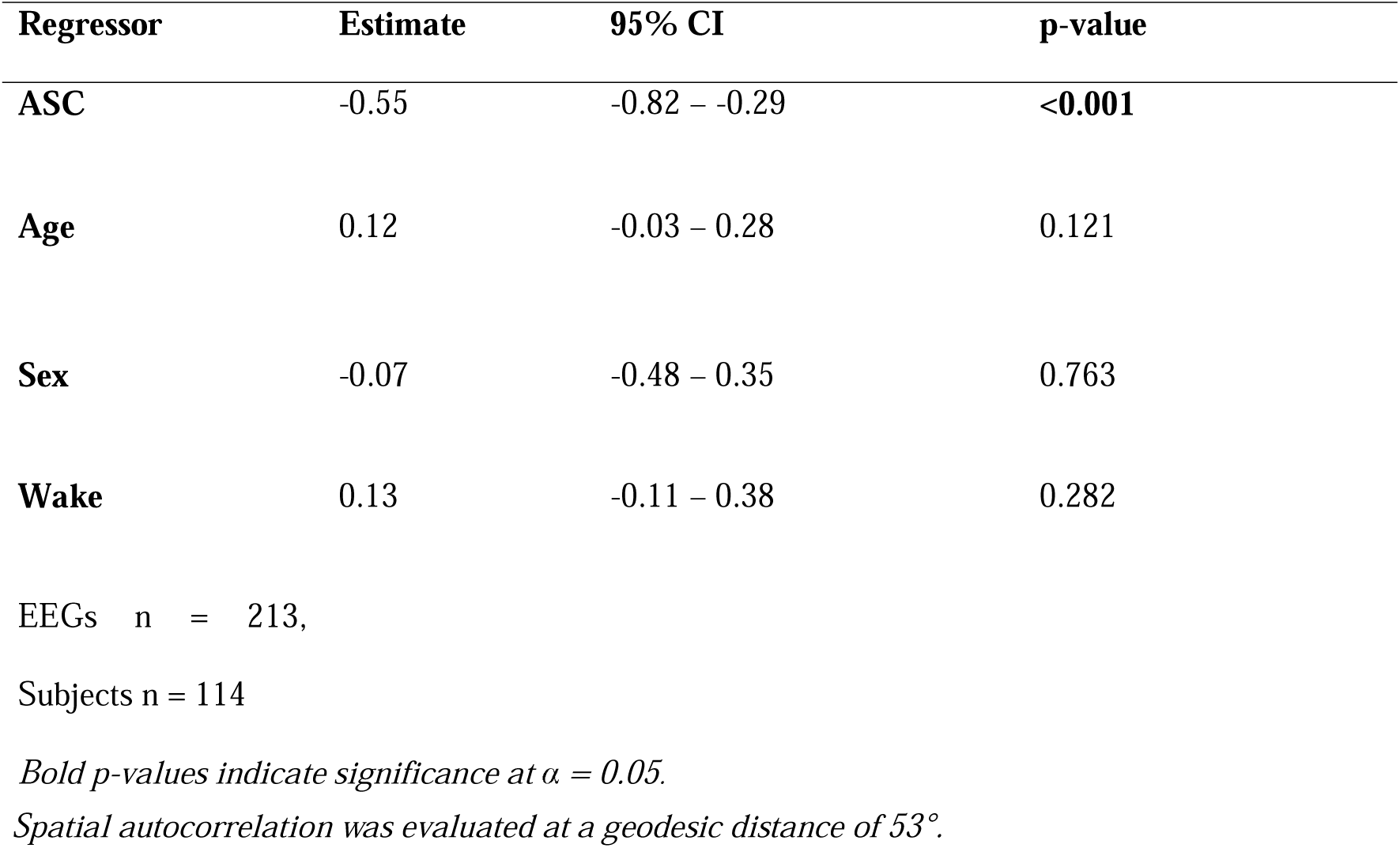
Aperiodic exponent spatial autocorrelation excluding participants who received melatonin as a sleep aid for their sleep EEG (discovery sample).

**Extended Data Table 5.**
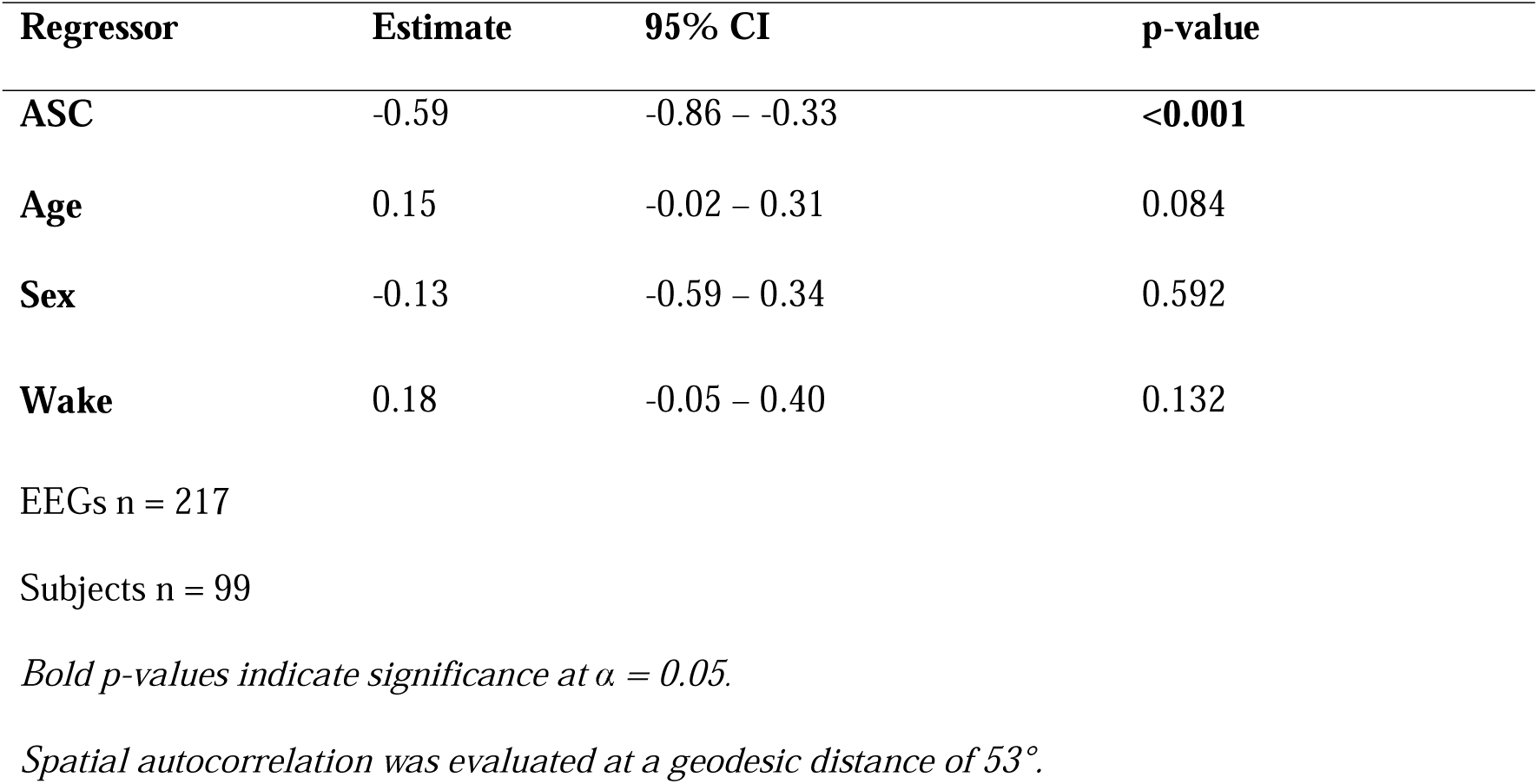
Aperiodic exponent spatial autocorrelation excluding ASC participants with intellectual disability/global developmental delay (discovery sample).

| **Regressor** | **Estimate** | **95% CI** | **p-value** |
| --- | --- | --- | --- |
| **ASC** | -0.59 | -0.86 – -0.33 | **<0.001** |
| **Age** | 0.15 | -0.02 – 0.31 | 0.084 |
| **Sex** | -0.13 | -0.59 – 0.34 | 0.592 |
| **Wake** | 0.18 | -0.05 – 0.40 | 0.132 |

EEGs n = 217
Subjects n = 99
*Bold p-values indicate significance at $\alpha = 0.05$ .*
*Spatial autocorrelation was evaluated at a geodesic distance of 53°.*

**Extended Data Table 6.** Aperiodic exponent spatial autocorrelation excluding ASC participants with epileptiform or slow wave anomalies (discovery sample).

| **Regressor** | **Estimate** | **95% CI** | **p-value** |
| --- | --- | --- | --- |
| **ASC** | -0.70 | -0.95 – -0.45 | **<0.001** |
| **Age** | 0.17 | 0.04 – 0.31 | **0.011** |
| **Sex** | -0.10 | -0.42 – 0.21 | 0.530 |
| **Wake** | 0.05 | -0.03 – 0.39 | 0.644 |

EEGs n = 232
Subjects n = 111
*Bold p-values indicate significance at $\alpha = 0.05$ .*
*Spatial autocorrelation was evaluated at a geodesic distance of 53°.*

**Extended Data Table 7a.** Aperiodic exponent spatial autocorrelation in age and sex matched samples from the HBN dataset.

| Regressor | Estimate | 95% CI | p-value |
| --- | --- | --- | --- |
| **ASC** | -0.43 | -0.79 – -0.06 | **0.022** |
| **Age** | 0.04 | -0.01 – 0.09 | 0.09 |
| **Sex** | 0.12 | -0.35 – 0.59 | 0.62 |

EEGs n = 832

**Extended Data Table 7b.** Aperiodic exponent spatial autocorrelation adjusting for coexisting conditions in the HBN dataset.

| Regressor | Estimate | 95% CI | p-value |
| --- | --- | --- | --- |
| **ASC** | -0.35 | -0.63 – -0.07 | **0.014** |
| **ADHD** | 0.03 | -0.18 – 0.24 | 0.778 |
| **Intellectual Disability** | 0.16 | -0.48 – 0.81 | 0.619 |
| **Age** | 0.25 | 0.15 – 0.35 | **<0.001** |
| **Sex** | -0.18 | -0.40 – 0.04 | 0.107 |

EEGs n = 2587
*Bold p-values indicate significance at $\alpha = 0.05$ .*
*Spatial autocorrelation was evaluated at a geodesic distance of 53°.*

**Extended Data Table 8a.** Aperiodic exponent spatial autocorrelation in the NIH dataset adjusting for eyes closure (mixed-effects model).

| **Regressor** | **Estimate** | **95% CI** | **p-value** |
| --- | --- | --- | --- |
| ASC (AD) | -0.18 | -0.33 – -0.03 | **0.020** |
| Sex (M) | 0.07 | -0.07 – 0.21 | 0.343 |
| Age | 0.09 | 0.02 – 0.16 | **0.013** |
| Eyes (open) | -0.46 | -0.60 – -0.31 | **<0.001** |

EEGs n = 1396

**Extended Data Table 8b.** Aperiodic exponent spatial autocorrelation in the NIH dataset in age and sex matched samples.

| **Regressor** | **Estimate** | **95% CI** | **p-value** |
| --- | --- | --- | --- |
| ASC (AD) | -0.19 | -0.36 – -0.02 | **0.044** |
| Sex (M) | 0.29 | 0.09 – 0.49 | **0.004** |
| Age | 0.02 | -0.07 – 0.11 | 0.672 |

EEGs n = 702

**Extended Data Table 8c.** Aperiodic exponent spatial autocorrelation in the NIH dataset (including the lower penetrance category “ASD” from Dede et al.).

| **Regressor** | **Estimate** | **95% CI** | **p-value** |
| --- | --- | --- | --- |
| ASC (AD + ASD) | -0.19 | -0.35 – -0.02 | **0.025** |
| Sex (M) | 0.10 | -0.06 – 0.27 | 0.208 |
| Age | 0.14 | 0.06 - 0.22 | **0.001** |

EEGs n = 1038

**Extended Data Table 8d.** Aperiodic exponent spatial autocorrelation in the NIH dataset (including the lower penetrance category “ASD” from Dede et al.) in age and sex matched samples.

| **Regressor** | **Estimate** | **95% CI** | **p-value** |
| --- | --- | --- | --- |
| ASC (ASD + AD) | -0.15 | -0.32 – 0.02 | 0.081 |
| Sex (M) | 0.22 | 0.05 – 0.40 | **0.013** |
| Age | 0.12 | 0.03 – 0.20 | **0.007** |

EEGs n = 834
*Bold p-values indicate significance at $\alpha = 0.05$ .*
*Spatial autocorrelation was evaluated at a geodesic distance of 53°.*

**Extended Data Table 9.** Nested logistic regression model comparison for ASC classification.

|  | **Discovery (19ch, n = 235)** |  | **HBN (128ch, n = 2587)** |  |
| --- | --- | --- | --- | --- |
| **Model** | **AIC** | **LR $\chi^2$ vs A** | **AIC** | **LR $\chi^2$ vs A** |
| A: age + sex | 326.8 | – | 2212.5 | – |
| B: A + mean exponent | 323.8 | 5.00 (p = 0.025) | 2209.7 | 4.83 (p = 0.028) |
| C: A + SD exponent | 328.8 | 0.00 (p = 0.950) | 2214.5 | 0.01 (p = 0.910) |
| D: A + mesoscale zI | **317.3** | 11.53 (p < 0.001) | 2208.5 | 6.00 (p = 0.014) |
| E: A + mean + SD + zI | 318.0 | – | **2206.9** | – |

| **Contrast** | **Discovery** | **HBN** |
| --- | --- | --- |
| E vs B (spatial measures add to mean?) | $\chi^2(2) = 9.84, \mathbf{p = 0.007}$ | $\chi^2(2) = 6.80, \mathbf{p = 0.03}$ |
| E vs D (level measures add to zI?) | $\chi^2(2) = 3.31, p = 0.19$ | $\chi^2(2) = 5.63, p = 0.06$ |

*Best AIC per cohort in bold. Bold p-values indicate significance at $\alpha = 0.05$ .*

**Extended Data Table 10.** E/I spatial autocorrelation and ASC traits penetrance in the discovery sample.

| **Regressor** | **Estimate** | **95% CI** | **p-value** |
| --- | --- | --- | --- |
| **ADOS CSS** | 0.00 | -0.16 – 0.16 | 0.990 |
| **Age** | 0.26 | -0.01 – 0.53 | 0.061 |
| **Sex** | -0.01 | -0.40 – 0.39 | 0.976 |
| **Wake** | 0.31 | 0.04 – 0.59 | **0.026** |

EEGs n = 184
Subjects n = 102
*Bold p-values indicate significance at $\alpha = 0.05$ .*
*Spatial autocorrelation was evaluated at a geodesic distance of 53°.*

**Supplementary Table 1.** Peak Alpha Frequency spatial autocorrelation.

| *Dataset* | **Regressor** | **Estimate** | **95% CI** | **p-value** |
| --- | --- | --- | --- | --- |
| **Discovery** |  |  |  |  |
| **sample**<br>**n = 204** | ASC | 0.01 | -0.29 – 0.31 | 0.948 |
|  | Sex (M) | -0.16 | -0.54 – 0.21 | 0.387 |
|  | Age | -0.03 | -0.18 – 0.12 | 0.676 |
| **HBN** |  |  |  |  |
| **n = 2545** | ASC | -0.70 | -1.10 – -0.30 | **0.001** |
|  | Sex (M) | 0.07 | -0.23 – 0.38 | 0.630 |
|  | Age | 0.11 | 0.07 – 0.15 | **<0.001** |
| **NIH** |  |  |  |  |
| **n = 932** | ASC | 0.05 | -0.11 – 0.21 | 0.544 |
|  | Sex (M) | -0.04 | -0.18 – 0.13 | 0.746 |
|  | Age | 0.02 | -0.06 – 0.10 | 0.605 |

*Bold p-values indicate significance at $\alpha = 0.05$ .*
*Spatial autocorrelation was evaluated at a geodesic distance of 53°.*

**Supplementary Table 2.** Aperiodic offset spatial autocorrelation.

| *Dataset* | **Regressors** | **Estimates** | **95% CI** | **p-value** |
| --- | --- | --- | --- | --- |
| **Discovery** |  |  |  |  |
| **sample**<br>**n = 235** | ASC | -0.65 | -0.95 – -0.34 | **<0.001** |
|  | Sex (M) | -0.08 | -0.44 – 0.29 | 0.679 |
|  | Age | 0.05 | -0.10 – 0.20 | 0.511 |
| **HBN** |  |  |  |  |
| **n = 2587** | ASC | -0.02 | -0.36 – 0.31 | 0.882 |
|  | Sex (M) | 0.33 | 0.07 – 0.59 | **0.011** |
|  | Age | 0.15 | 0.11 – 0.18 | **<0.001** |

*Bold p-values indicate significance at $\alpha = 0.05$ .*
*Spatial autocorrelation was evaluated at a geodesic distance of 53°.*

## Notes

### Clinical Protocols

https://osf.io/4q83m

### Funding Statement

This study did not receive any funding.

### Author Declarations

All the procedures were approved by the local ethics committee (Comitato Etico Territoriale Interaziendale AOU Città della Salute e della Scienza di Torino; previously Comitato Etico Città della Salute e della scienza di Torino) and parents or legal tutors of ASD patients gave informed consent. Control EEGs were collected anonymously.

### Summary of Updates

The previous version was framed as a biomarker study. It lacked the continuous spatial autocorrelation framework, the mesoscale specificity, the structural MRI integration, and the Healthy Brain Network large validation cohort.

